# Rural Emergency 360°: A mixed-methods analysis of barriers and priorities for equitable emergency care in Québec

**DOI:** 10.64898/2026.02.02.26345381

**Authors:** Richard P. Fleet, Catherine Turgeon-Pelchat, Fatoumata Korika Tounkara, Gilles Dupuis, Jean-Paul Fortin, Jocelyn Gravel, Mathieu Ouimet, Julie Théberge, France Légaré, Hassane Alami

## Abstract

**Background:** Rural emergency departments (EDs) are critical to ensuring equitable access to acute care, yet face persistent systemic challenges. In Québec, Canada, reforms to healthcare governance, funding and resource allocation, and service delivery have transformed rural ED operations. This study aimed to document characteristics, challenges, and improvement priorities for all rural EDs in the province.

**Methods:** A participatory mixed-methods design was used. 26 rural EDs in Québec were included. Data sources comprised administrative statistics, structured site surveys, individual stakeholder semi structured interviews, and a validation survey of identified local champions. Analyses comprised a triangulation of the quantitative and qualitative data using transversal thematic analysis to determine common issues. Potential solutions identified were validated through stakeholder questionnaires. The study was reported in accordance with the COREQ reporting guideline.

**Results:** Most respondents were women (64%) and professionals with more than 5 years of experience. Four main themes were identified: governance, healthcare organization, access to resources, and professional practice. Governance challenges included reduced local autonomy, administrative complexity, and budgeting models poorly adapted to rural realities. Participants emphasized the need for standardized but locally flexible administrative processes, regional emergency service managers, and rural-sensitive performance metrics. Organizational barriers included geographic isolation, limited access to primary care, and difficulties with interfacility transfers due to referral-center capacity and ambulance shortages. Resource constraints centered on shortages of human resources, diagnostic services and specialty coverage, especially anesthesia, obstetrics, and psychiatry. Professional practice was shaped by the need to maintain broad competencies in low-volume contexts, while contending with professional isolation and proximity to patients. Local champions prioritized expanding telemedicine, strengthening prehospital services, enhancing continuing education, and implementing tailored recruitment strategies.

**Conclusion:** This study provides the first province-wide documentation of characteristics, challenges, and improvement priorities for all rural EDs. Findings highlight the need for systemic reforms that restore local decision-making authority, strengthen transfer and prehospital capacity, expand telehealth and specialty access, and support professional development. These results provide a foundation for evidence-based policies and actions to sustain equitable emergency care in rural regions.

## Introduction

Emergency services in rural areas serve as a vital safety net for populations that are older, in poorer health, and at greater risk of trauma [1–4]. In Canada, over 300 rural Emergency Departments (EDs) handle approximately 20,000 visits per year on average, with around 25% of these cases involving seriously ill patients, many of whom require inter-facility transfers [5,6]. These rural EDs operate within a challenging organizational framework defined by limited alternative care options, a high proportion of patients lacking regular healthcare providers, and persistent difficulties in recruiting and retaining physicians [7]. Despite these challenges, rural emergency services remain critically under-researched, with few evidence-based standards or guidelines tailored to their unique context, despite longstanding calls for such frameworks [3,5,8].

Research from Québec demonstrates that clinical outcomes in rural settings are significantly poorer compared to urban areas [9,10]. Patients in these rural regions are three times more likely to die from trauma compared to their urban counterparts [9], and face a 25% higher mortality rate following a stroke [10]. These disparities align with both national and international studies, emphasizing the urgency of improving rural healthcare systems [10].

Québec’s publicly funded healthcare system is organized into large regional health and social services centers known as Centres intégrés de santé et de services sociaux (CISSS) and Centres intégrés universitaires de santé et de services sociaux (CIUSSS), established as part of major governance reforms in 2015 (see Alami et al. [11] for more detail on the Québec public health system). The CISSS and CIUSSS are responsible for overseeing hospitals, long-term care, public health, and community services within their regions. Québec includes approximately 117 hospitals, including 26 designated rural EDs. While public administration facilitates standardization, the centralization of governance has significantly diminished local decision-making capacity, particularly in rural areas, further complicating care delivery. However, we still know little about the impact of this reorganization of the system.

Against this backdrop, this study employs a Participatory Action Research (PAR) framework across all 26 rural EDs in Québec to address the lack of contextualized evidence[12,13]. Thus we sought to: 1) provide a comprehensive overview of the unique challenges facing rural emergency services from diverse stakeholder perspectives; 2) identify feasible and prioritized solutions to enhance quality and performance; and 3) examine barriers and facilitators affecting implementation and sustainability. This will in turn provide the needed guidance in applying evidence-based recommendations for meaningful, locally-adapted improvements in emergency care in the future.

### Relation to previous work

This article presents, for the first time, the complete scientific report that served as one of three knowledge translation (KT) strategies evaluated in a previously published *PLOS One* study (“Through the big top: An exploratory study of circus-based artistic knowledge translation in rural healthcare services, Québec, Canada.” [14]). Led by Théberge and colleagues, this earlier study compared a circus performance, a scientific PowerPoint presentation (webinar), and a written research report to determine their relative effectiveness in disseminating rural health research findings as part of an arts-based knowledge translation (ABKT) intervention. The present article does not duplicate results from the earlier publication; rather, it offers a full, standalone version of the written research report that was previously summarized for evaluation purposes. In doing so, it provides readers with direct access to the empirical findings that underpinned the ABKT intervention. This ensures transparency, reproducibility, and broader accessibility while enabling critical reflection and comparison between traditional and artistic dissemination formats.

## Methods

### Study design

This study applied a PAR approach to promote active stakeholder engagement and support the practical adoption of findings within participating settings [15,16]. Participants were informed that PAR emphasizes co-construction, fostering collaboration between researchers and participants to facilitate the successful implementation of solutions[16]. A mixed-methods design, incorporating both qualitative and quantitative components, was selected to offer comprehensive and rigorous research objectives while maintaining methodological rigor and adaptability through integrated qualitative and quantitative components[17]. The reporting guidelines COREQ was used. Ethical approval was obtained from the Research Ethics Committee of CISSS Chaudière-Appalaches (approval number: MP-23-2017-396).

### Participating emergency departments

All eligible rural EDs in Québec that met the study’s inclusion and exclusion criteria were included (Fig 1). The eligibility criteria aimed to ensure relative homogeneity across sites and support meaningful comparisons. EDs were eligible if they: 1) provided 24/7 medical coverage in hospitals with inpatient beds; 2) were located in municipalities with fewer than 15,000 residents according to the 2016 Canadian census [16]; and 3) were situated more than 50 minutes away from a secondary or tertiary trauma center [18]. Due to logistical constraints, EDs located in the Nunavik and Terres-Cries-de-la-Baie-James regions were excluded.

**Fig 1.**
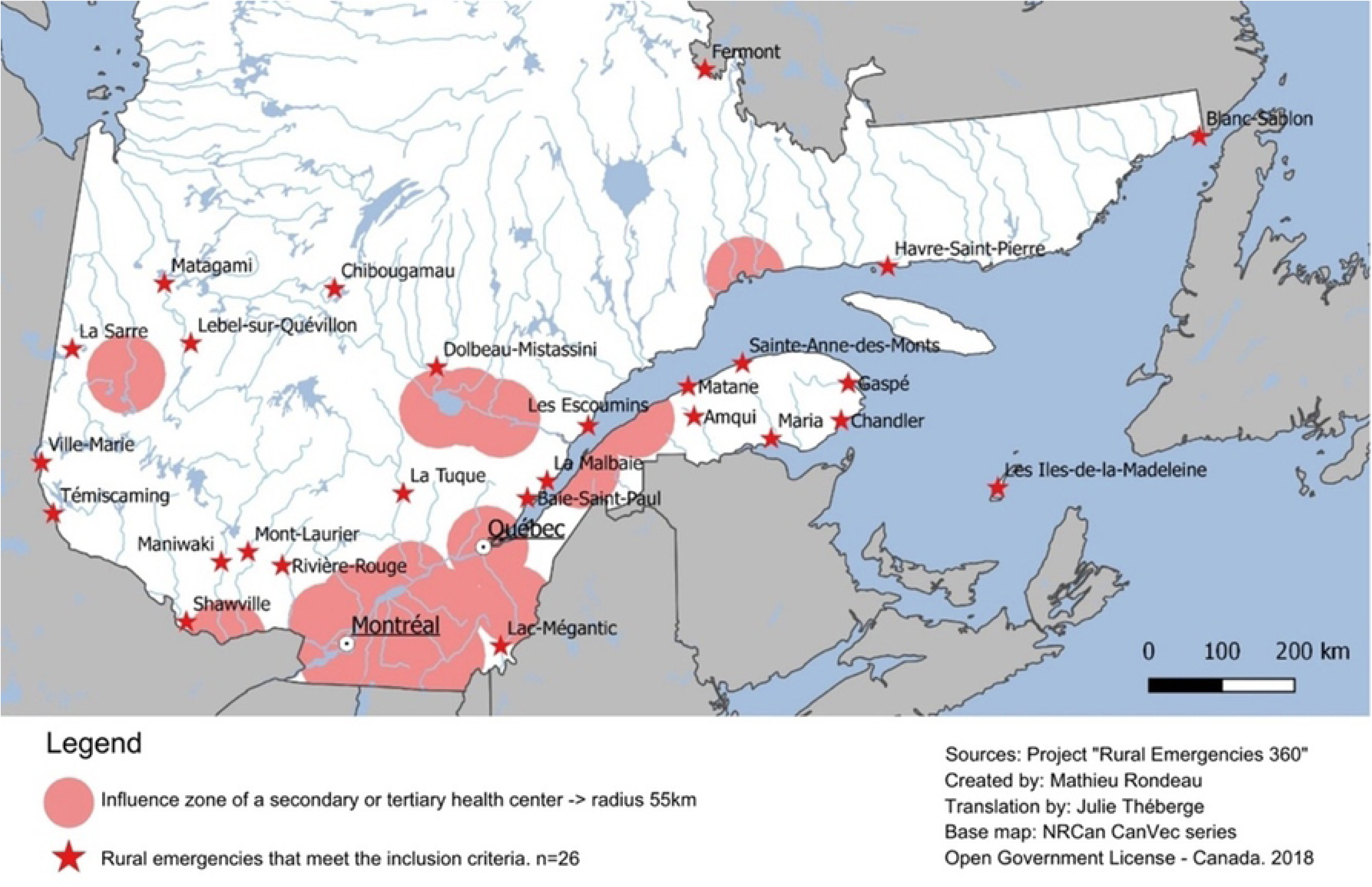
Map of rural EDs participating in the Rural Emergencies 360 project.

Two pilot studies were conducted in 2015 and 2016 to refine the study methodology and data collection tools. These pilots were implemented in three rural EDs that also participated in the Rural Emergency 360 project. A total of 91 participants with diverse profiles took part in these studies through individual and group interviews. Findings from these pilots have been reported elsewhere [13,19], and are not repeated here, except where directly relevant.

### Recruitment

Recruitment was guided by two principles: 1) ensuring diverse viewpoints; and 2) achieving data saturation defined as “…achieving data saturation defined as the point where further interviews did not generate new perspectives on rural emergency care processes, challenges, or solutions.”. Eight participant categories and three ED types were identified a priori (Fig 2). The recruitment process was designed to capture all categories across ED types and regions. Participants were primarily recruited through local champions who suggested potential candidates for each category. The research team subsequently contacted these individuals directly. Additional participants were identified via snowball sampling and direct outreach to individuals with key roles. Data collection continued until all participant categories and ED types were represented and no new themes emerged [20,21].

**Fig 2.**
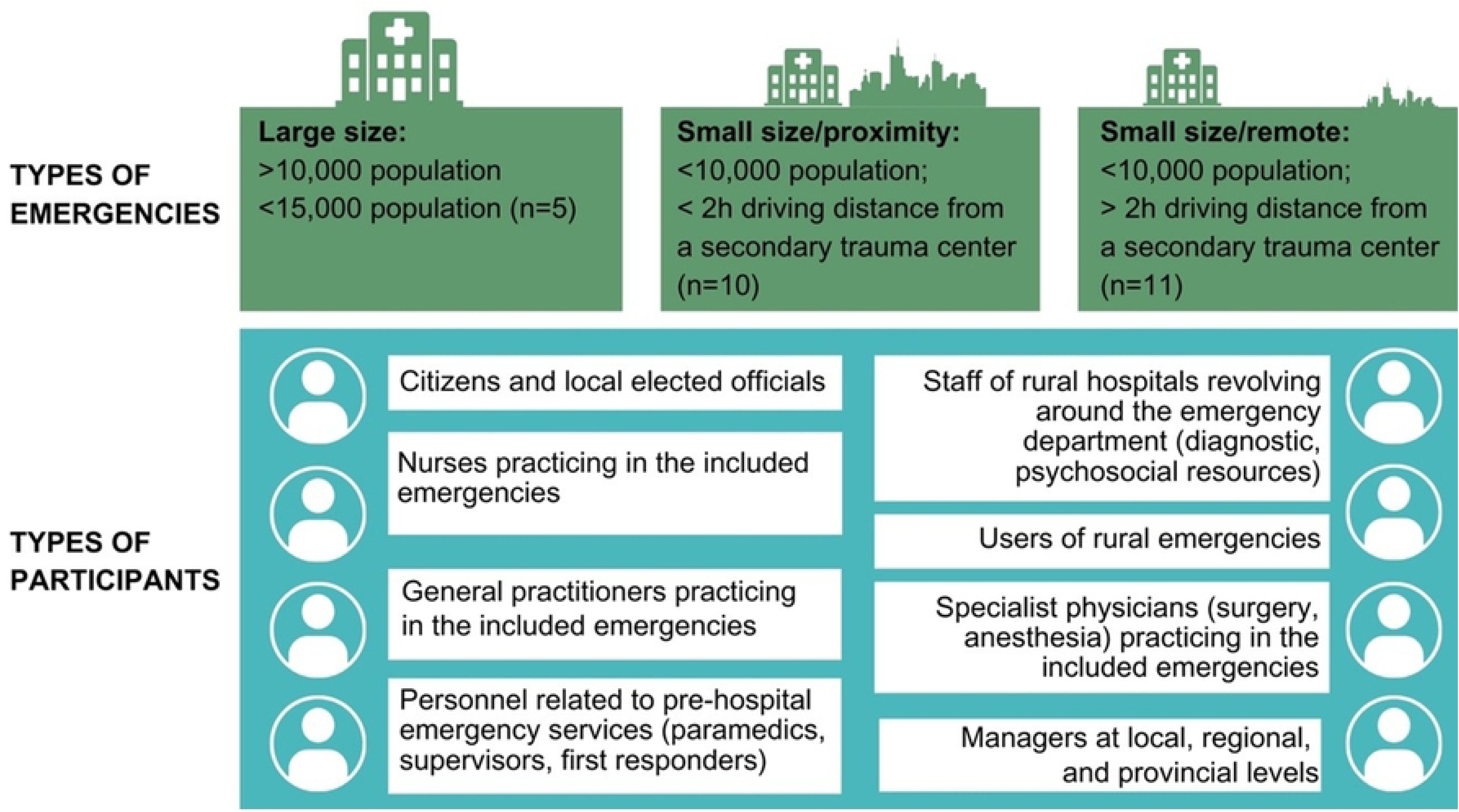
Division of participating EDs into three types and identification of participant types.

## Data collection

### Quantitative component

The quantitative component of this study aimed to provide a detailed statistical profile of Québec’s rural EDs including wait times, staffing, and available resources. Data were drawn from the following three sources: the Québec Ministère de la Santé et des Services sociaux (MSSS), Directorate of Hospital Services, Prehospital and Emergency Services (2016–2017 emergency data); annual statistical reports (2016–2017) from participating institutions; and a structured questionnaire completed by a designated “champion” or representative from each participating ED. Data covering the year 2016–2017 were first analyzed using descriptive statistics in Microsoft Excel (Microsoft Corporation) and subsequently validated by local champions. Corrections provided during validation were incorporated into the final dataset.

### Qualitative component

The qualitative component was designed to: 1) engage stakeholders including decision-makers, professionals, and citizens in PAR; 2) provide a comprehensive analysis of challenges in rural EDs from multiple perspectives; 3) identify locally developed or proposed solutions to improve quality and performance; and 4) explore barriers and facilitators influencing the implementation of these solutions.

### Interviews

Semi-structured individual and group interviews were conducted between September 2017 and July 2018 by two research professionals. Interviews were carried out in person, via videoconference, or by telephone using guides tailored to participant type and format[12]. All interviews were recorded and transcribed verbatim. Participants were informed about the study details, provided written informed consent and completed a socio-demographic questionnaire. Interviews and focus groups were conducted by <u>Catherine Turgeon-Pelchat</u> (Master’s in Anthropology and study coordinator with 10 years of experience in qualitative research in rural healthcare).

### Data analysis

Interview data were analyzed using an inductive thematic approach in NVivo, allowing themes to emerge naturally from the data (QSR International Pty Ltd). Coding began inductively to identify emerging issues and solutions, which were then organized into themes and subthemes[12,13]. This process also incorporated insights from earlier pilot studies [13,19], with adaptations to reflect the new dataset. Three research professionals conducted the coding collaboratively to ensure consistency and clarity in categorization. Results were refined through discussions with co-researchers and a qualitative methods expert. The cross-sectional analysis approach allowed for comparisons to be made across diverse settings in order to identify common challenges and potential solutions for rural EDs.

### Local validation of qualitative data

To validate findings from the qualitative data, a survey was developed and piloted by members of the research team. The survey was distributed via LimeSurvey (GmbH, Hamburg, Germany). The survey summarized key results and asked respondents to confirm their accuracy (e.g., “Does this description reflect your environment?”). It also included prioritization questions such as: “Do you consider implementing this solution a priority?”. Survey responses were used to triangulate findings and to assess the alignment of proposed solutions with the priorities of stakeholders. The results of this validation exercise are presented in the Results section.

## Results

### Research participants

All 26 EDs participated in the quantitative component. Table 1 presents the distribution of participants across categories and interview formats. Among the 26 designated champions, 11 (42%) also took part in interviews, and 19 (73%) completed the questionnaire; nine participants (35%) contributed to both components of the study. Three interview participants were involved exclusively in pilot studies. Overall, every ED engaged in at least one component, and 77% participated in two or more study components. In total, 83 individuals participated in 58 individual or group interviews (duration of 60 to 90min per interview). Key socio-demographic characteristics of interviewed participants are summarized in Table 2. Most respondents were women (64%) and were mid- to late-career professionals (i.e., having more than 5 years of experience).

**Table 1.** Types of participants in the individual and group interviews.

| Participants | Individual<br>interviews<br>(n = 48) | Group<br>interviews<br>(n = 10) | Total<br>(n = 58) |
| --- | --- | --- | --- |
| Physicians (general practitioners and specialists) | 15 (31%) | 0 (0%) | 15 (26%) |
| Managers (MHSS, CISSS/CIUSSS, ED) | 11 (23%) | 1 (10%) | 12 (21%) |
| Nursing staff (ED, all shifts) | 8 (17%) | 2 (20%) | 10 (17%) |
| Citizens (elected officials, concerned citizens, users) | 6 (13%) | 3 (30%) | 9 (15%) |
| Psychosocial staff (social workers, psychologists) | 5 (10%) | 0 (0%) | 5 (9%) |
| Diagnostic services (radiology, lab, ultrasound, etc.) | 2 (4%) | 2 (20%) | 4 (7%) |
| Prehospital emergency services (paramedics, supervisors, first responders) | 1 (2%) | 2 (20%) | 3 (5%) |

MHSS = Ministry of Health and Social Services; CISSS = Centres intégrés de santé et de services sociaux; CIUSSS = Centres intégrés universitaires de santé et de services sociaux (CIUSSS); ED = emergency department

**Table 2.** Key sociodemographic characteristics of interview participants.

| Characteristic | Participants (n = 83) |
| --- | --- |
| Sex |  |
| Male | 30 (36.1%) |
| Female | 53 (63.9%) |
| Age (y), mean ± SD (n = 69) | 45.7 ± 57.2 |
| Experience in current position (y), mean ± SD (n = 56) | 12.8 ± 24.3 |
| Experience at hospital/town studied (y), mean ± SD (n = 50) | 13.3 ± 23.7 |

### Emergency department characteristics

The 26 rural EDs studied are located in hospitals with an average of 31 ± 17 inpatient beds (excluding ED beds). Annual ED visits averaged 15,485 ± 5,276, of which most patients were triaged at level 4 or level 5 (79.6%; 12,498 visits). On average, 1,449 ± 1,124 patients (9.4%) left without being seen. Table 3 provides a detailed overview of the rural EDs participating in the study.

**Table 3.**
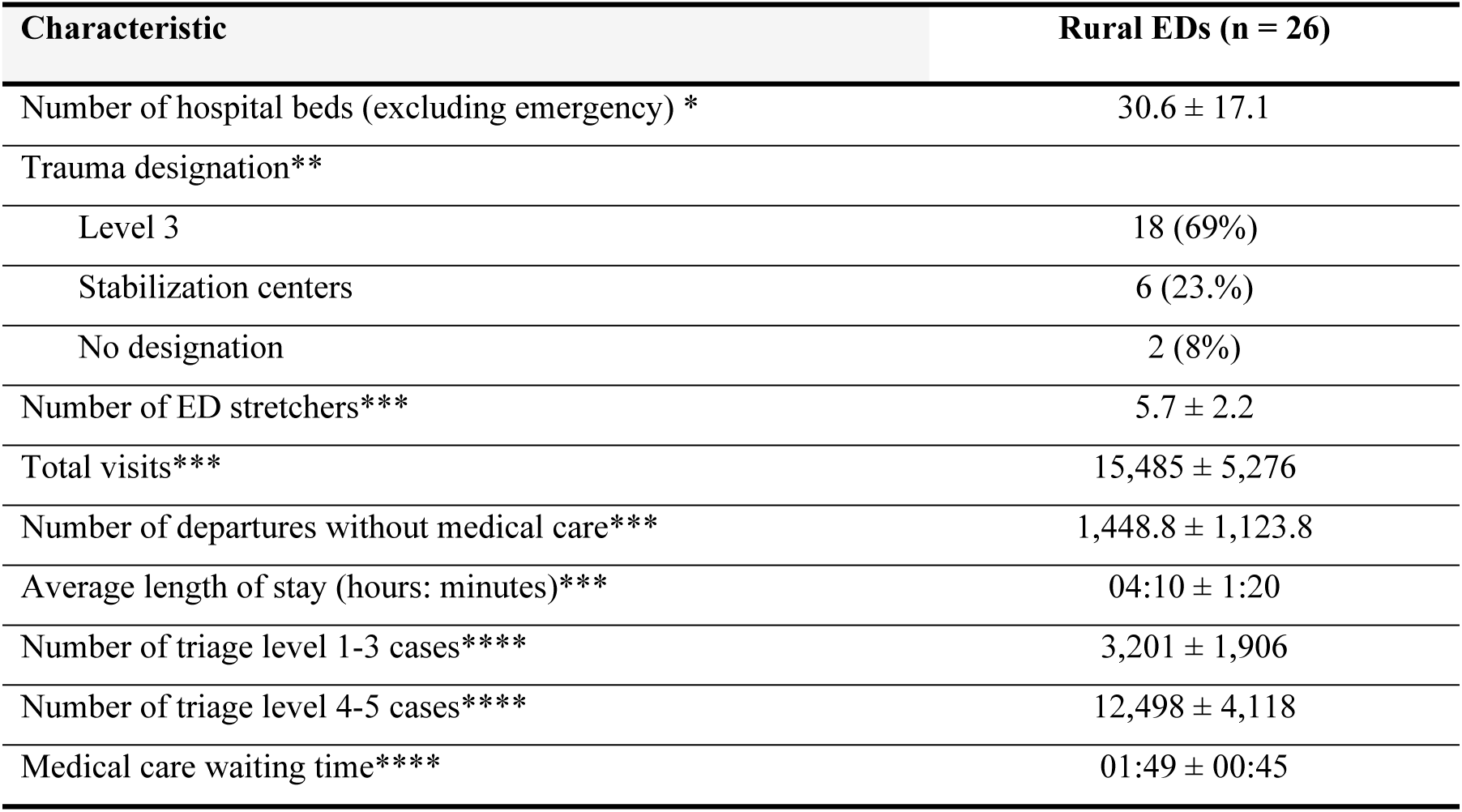
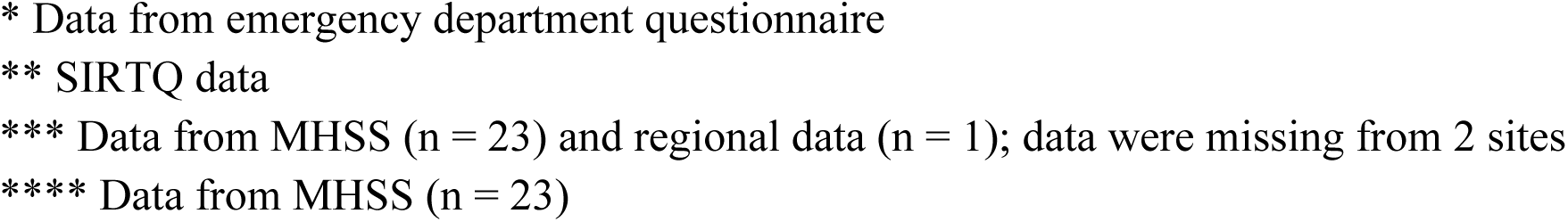
Characteristics of participating rural EDs.

| Characteristic | Rural EDs (n = 26) |
| --- | --- |
| Number of hospital beds (excluding emergency) * | 30.6 ± 17.1 |
| Trauma designation** |  |
| Level 3 | 18 (69%) |
| Stabilization centers | 6 (23.%) |
| No designation | 2 (8%) |
| Number of ED stretchers*** | 5.7 ± 2.2 |
| Total visits*** | 15,485 ± 5,276 |
| Number of departures without medical care*** | 1,448.8 ± 1,123.8 |
| Average length of stay (hours: minutes)*** | 04:10 ± 1:20 |
| Number of triage level 1-3 cases**** | 3,201 ± 1,906 |
| Number of triage level 4-5 cases**** | 12,498 ± 4,118 |
| Medical care waiting time**** | 01:49 ± 00:45 |

\* Data from emergency department questionnaire \*\* SIRTQ data \*\*\* Data from MHSS (n = 23) and regional data (n = 1); data were missing from 2 sites \*\*\*\* Data from MHSS (n = 23)

### Identification of challenges and opportunities for service enhancement

Findings from the quantitative questionnaire, qualitative interviews, and validation survey are presented together, organized by key themes to facilitate integration/triangulation across sources. This approach identifies major challenges and priority areas for improving rural emergency care. Fig 3 provides an overview of themes, highlighting links between rural-specific issues and actionable goals. The sections that follow present results by four main themes that were identified, along with relevant sub-themes: 1) governance; 2) healthcare organization; 3) access to resources; and 4) professional practice. In addition to the participant quotations highlighted in these sections, the complete and extended quotations from interview participants are available in the Supplementary Information (S1 Appendix) and have been similarly categorized into four main themes and related sub-themes.

**Fig 3.**
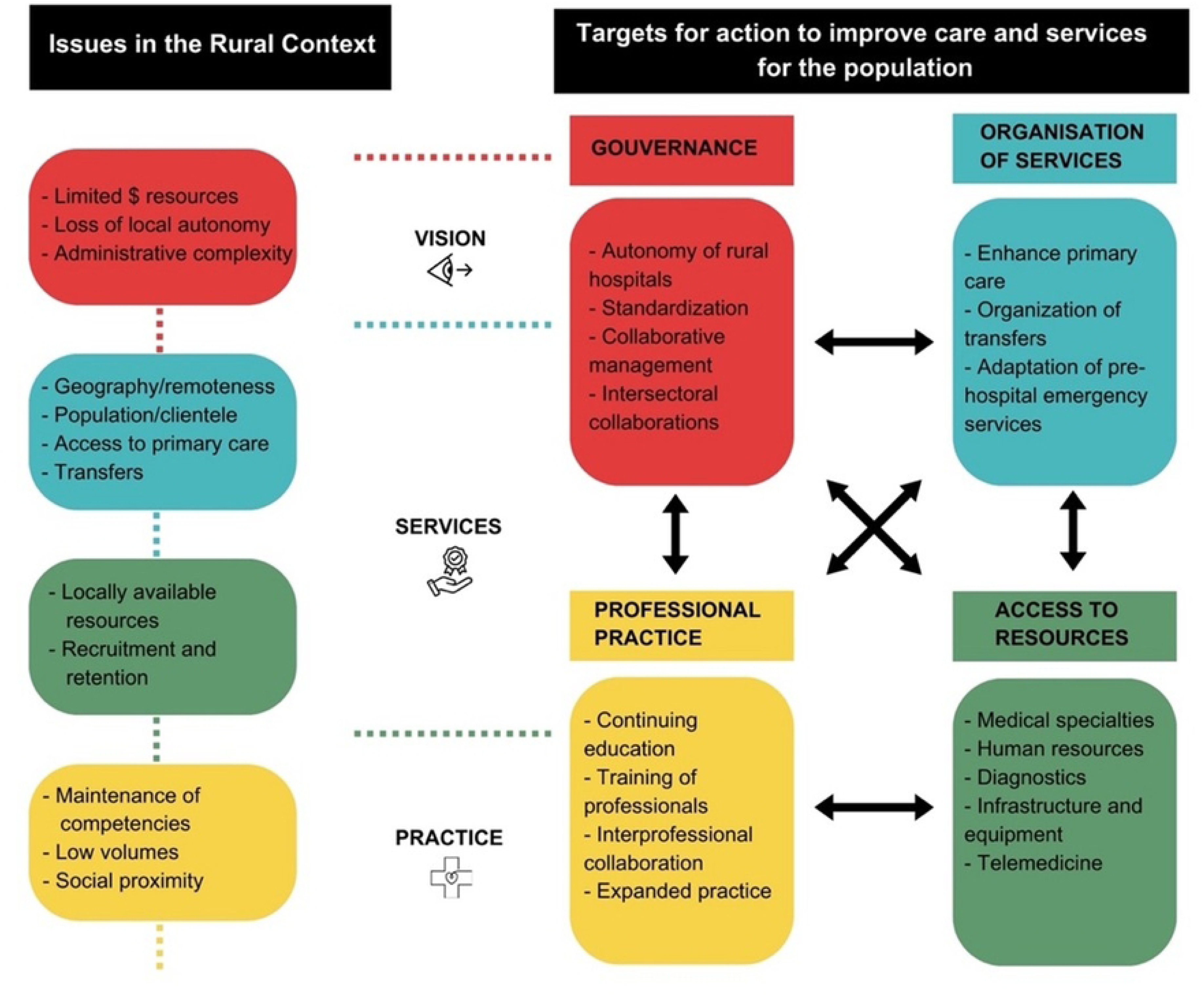
Issues and targets for service improvement.

### Theme 1: Governance

Governance emerged as a cross-cutting theme, encompassing the organizational, administrative, and political dimensions of rural emergency care. Participants highlighted three key challenges related to governance: financial constraints, limited local autonomy, and administrative complexity.

### Financial resources and funding models

Participants were quick to note the limited availability of financial resources and the impact this has on decision-making. Some participants noted that reliance on volumes to justify resource allocation contributed to the gradual erosion of rural services. Several participants felt that activity-based funding disadvantaged rural EDs with smaller patient volumes despite significant health needs:

> *We really have to approach it with a logic of “is it relevant? Is it efficient? Is it really worth it?” [EI53_ges]*

### Loss of local autonomy

In 2015, sweeping reforms led to the centralization of Québec’s health system with the merger of over 182 health and social service facilities into 34 integrated centres (CISSS/CIUSSS) [24]. Participants reported feeling that this change was widely perceived at the local level to have resulted in the centralization of decision-making, leaving rural facilities without any real influence over the health needs and priorities in their communities:

> *[Participant 1] if decisions are made 7 hours away, well… something gets lost in translation, and between three chairs! [FG09_cit]*

### Administrative complexity

The creation of the 34 CISSS/CIUSSS in 2015 introduced new layers of bureaucracy for rural EDs to navigate, often hindering communication and slowing down the problem-solving process. Participants described increased reliance on formal procedures instead of the informal exchanges that once characterized smaller organizations:

> *So the mobilization of a team is still there, but as soon as we start moving up the ranks and end up at the CISSS level, there’s nothing feasible in the short term. [EI35_md]*

### Standardization versus local adaptation

Champions strongly supported harmonization of clinical and administrative protocols but emphasized the need for flexibility and adaptation to local contexts. In the validation survey (n = 19), 55% prioritized regional committees as coordinators of protocol harmonization, 31% preferred national committees, and 14% favored individual services. None of the champions rated harmonization as low priority. One manager summarized the balance needed:

> *We must be careful between standardization and harmonization. I believe in harmonization. If you tell me “in a few years we should move towards this, towards such a process,” that’s fine. That means all the changes you make will be in that direction. But I think we should move towards harmonization. [EI60_ges]*

Potential action targets related to the theme of governance as well as their degree of prioritization, are summarized in Fig 4. according to the champions who participated in the validation questionnaire. Alongside a desire for personalization and to “not lose local colors” or “dilute local issues in the machine,” the 19 champions who participated in the validation questionnaire expressed a strong need for standardization, sharing of clinical and administrative resources with the CISSS/CIUSSS, and metrics to evaluate progress. The majority of champions (55%) indicated that “protocol harmonization” should be primarily coordinated by regional committees. “National committees” were also viewed as important coordinators by 31% of respondents. “Each service”, including hospitals and departments, were identified by 14% of respondents respectively. Notably, none of the respondents (0%) considered “protocol harmonization” to be a low-priority issue. These results underscore a strong preference for coordination at broader systemic levels, particularly regional, rather than at individual service points. One champion noted:

> *Then we waste an insane amount of time every three years redoing the protocols, checking them. They should put them on the Internet and share them with the whole network. [EI02_md]*

**Fig 4.**
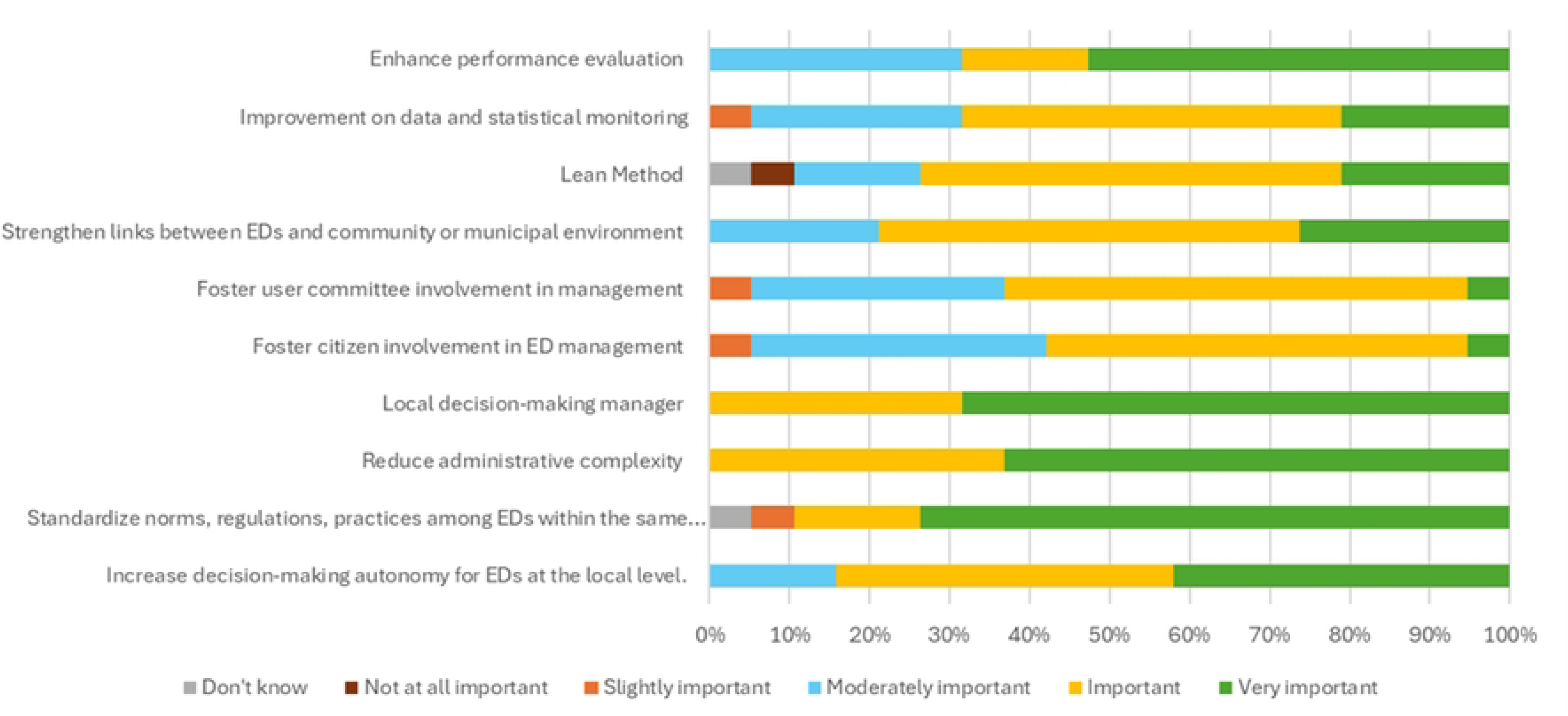
Responses (%) from champions (n = 19) to the following questions: “How much priority should be given to the following solutions to address challenges related to emergency management?”; and “How much priority should be given to the following solutions related to professional practice in rural settings?”.

### Performance monitoring

Performance metrics were described as a double-edged sword: they are useful for accountability and for sustaining bureaucratic processes, yet often poorly aligned with patient-centered care.

> *Of course, the demands from the ministry are enormous. I find that it is very focused on statistics, and not on the client, less on the clinical aspect: which we, the social workers, struggle a bit with because we are not stat machines, we help people! What they ask us to do for stats is not representative of a social worker’s job. [EI10_psy]*

Others described how meeting targets sometimes led to practices that undermined quality:

> *[Health minister] has given orders that patients must not stay more than 20 hours in the emergency room. They must not be… They must be moved when they are admitted. We have two hours to move the patient. If you don’t have a bed upstairs, you put them in the hallway to hide them. It’s awful, it’s terrible! [FG20_inf]*

### Collaborative and community-based governance bottom up approach

In some settings, collaborative governance (such as involving frontline staff, user committees, and physicians in leadership roles) was viewed as effective. Lean management and visual monitoring tools were also reported as helpful in linking frontline and administrative priorities:

> *We are even very advanced in terms of control rooms, coherence matrices, and visual stations deployed in the teams. I think that, at the administrative level, this is the link between senior management and field staff. [EI27_ges]*

Community mobilization and advocacy by citizens and elected officials were also credited with maintaining services such as dialysis or obstetrics:

> *Or often, it’s citizen pressure. I wouldn’t attribute the success of that solely to the leaders of the CISSS, because often it’s citizen pressure. [EI45_cit]*

### Theme 2: Organization of healthcare services

Participating rural EDs were located in small towns with a mean population of 5,905 ± 449 inhabitants (Source: Statistics Canada. Census 2016). These facilities are located at considerable distances from higher-level trauma centers (Table 5). Only two EDs were located less than 75 km from a level 1 or level 2 trauma center. Most facilities were located between 76 – 150 km (n = 9; 35%), and between 151 – 225 km (n = 6; 23%). There were five rural EDs (19%) located greater than 375 km (or absence of road) from a level 1 or 2 trauma center. The remoteness of most facilities from higher level trauma care frames the complexity of the organizational challenges facing rural EDs. Against this backdrop, participants highlighted three major issues: access to primary care, interfacility transfers, and prehospital emergency services.

**Table 4.** Proximity of rural EDs to the nearest level 1 or 2 trauma center.

| Proximity to Level 1 or Level 2 Trauma Centers* | Rural EDs (n = 26) |
| --- | --- |
| < 75 km | 2 (8%) |
| 76 - 150 km | 9 (35%) |
| 151 - 225 km | 6 (23%) |
| 226 - 300 km | 3 (11%) |
| 301 - 375 km | 1 (4%) |
| > 375 km (or absence of road) | 5 (19%) |

Mean population of 26 towns with participating rural EDs = 5,905 ± 4,491 (Source: Statistics Canada. Census 2016)
\* Source: Google Maps

**Table 5.** Access to various resources within the 26 participating facilities, 2016-2017.

| Resource | Rural EDs (n = 26) |
| --- | --- |
| Liaison nurse | 17 (65%) |
| Orderly | 14 (54%) |
| Telemedicine | 13 (50%) |
| Dialysis machine(s) | 10 (38%) |
| Designated medical coordinator (ER) | 8 (31%) |
| Heavy case/great user manager | 5 (19%) |

### Primary care and emergency department use

Champions prioritized strengthening access to primary care in the validation survey (Fig 5). Participants frequently linked ED crowding with limited access to primary care, noting that some non-urgent needs are managed in the ED due to the lack of alternatives:

> *As mentioned earlier, we have many patients who no longer have a family doctor. They come in for adjustments to their diabetes medications. That’s not my expertise. Sometimes we end up doing office work in the emergency department. [E161_md]*

**Fig 5.**
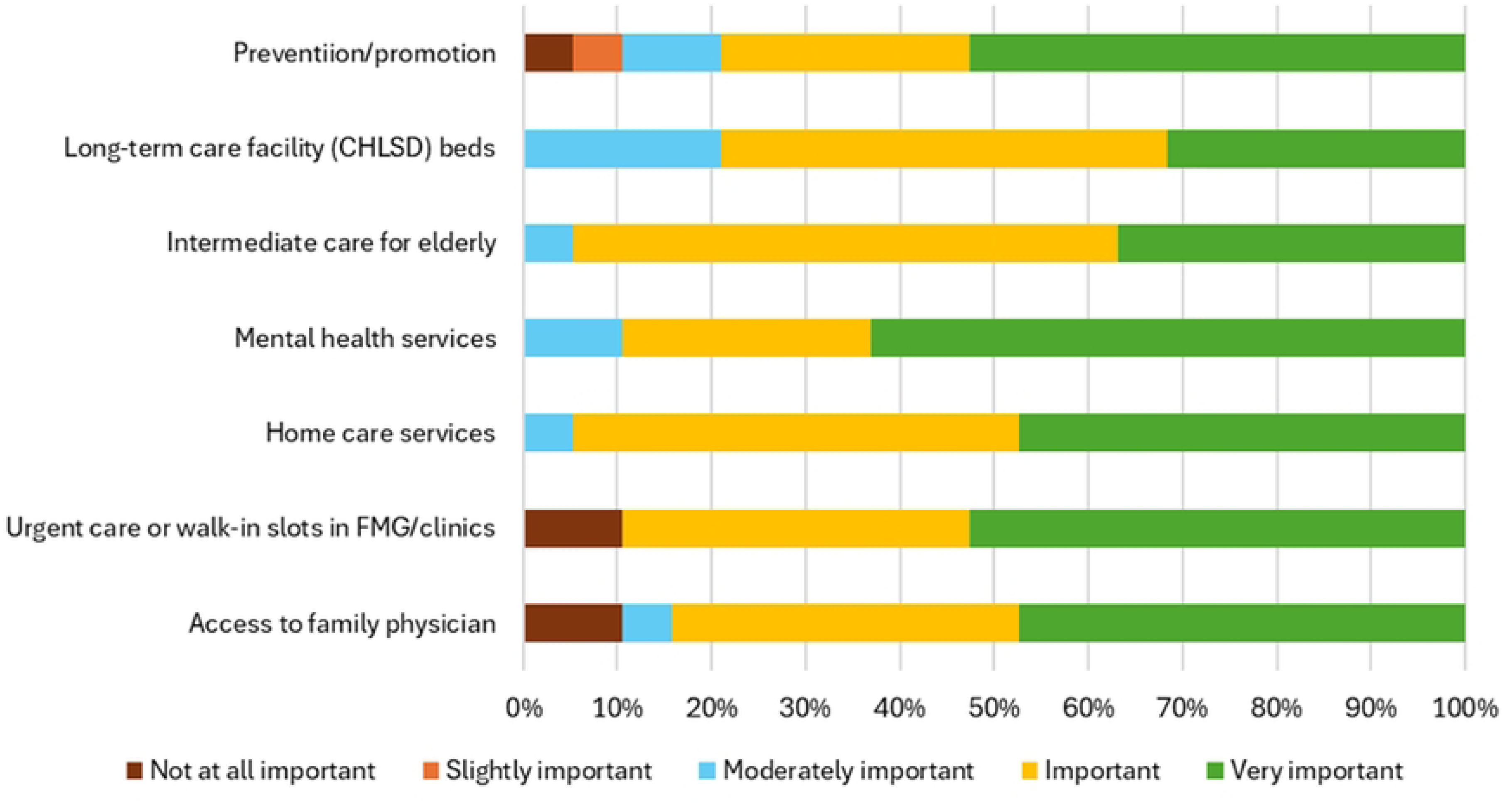
Responses (%) from champions (n = 19) to the question: “How much priority should be given to improving these aspects of access to primary care in your environment?” CHLSD = Centre d’hébergement et de soins de longue durée; FMG = Family Medicine Group.

### Interfacility transfers

Data from the ED questionnaire (n = 25) estimated a mean ± SD of 541 ± 933.3 interfacility transfers from EDs each year. The top five reasons provided for these interfacility transfers, as ranked by respondents (n = 26) in order of importance from most to least important were: 1) cardiology; 2) orthopedics; 3) trauma; 4) psychiatry; and 5) pediatrics. Champions ranked improvements in transfer coordination and safety among the highest priorities in the validation survey (Fig 6). Transfers to higher-level trauma and specialty centers were described as frequent, resource-intensive, and often delayed due to capacity shortages at receiving hospitals:

> *Often, they’ll tell me, ‘We don’t have a bed. We don’t have space.’ Then we have to shop around for a hospital. That’s really what it is! So it’s a waste of time for us, for emergency doctors, for nurses too. For the patient, it’s lost time where they could have already received their treatment or the required care that we don’t have here. [EI13_md]*

**Fig. 6.**
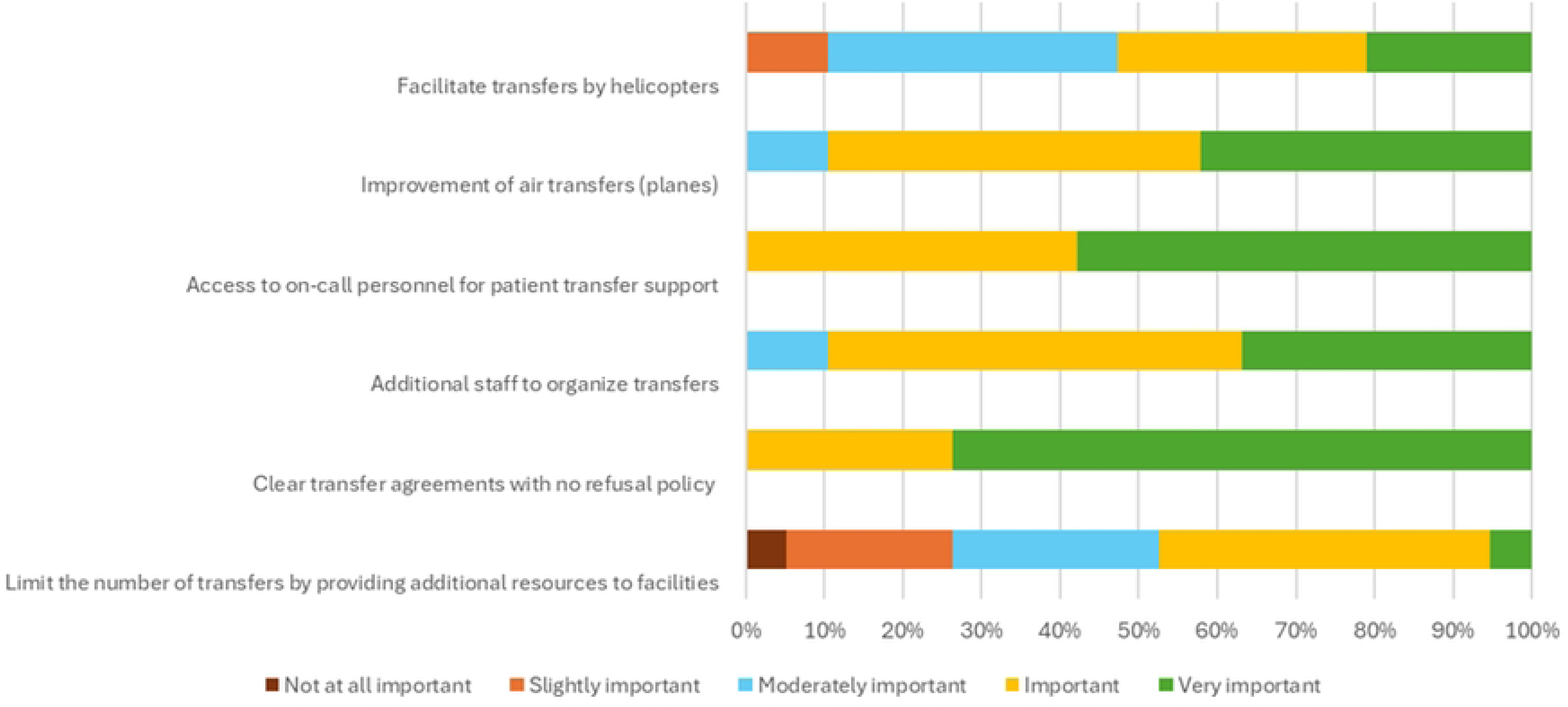
Responses (%) from champions (n = 19) to the question: “How much priority should be given to the following solutions to address challenges with interfacility transfers?”.

### Prehospital emergency services

Ambulance availability and long response times were highlighted as critical bottlenecks. Champions strongly endorsed strengthening prehospital capacity, including personnel and equipment, as an urgent need (Fig 7). When ambulances were diverted for interfacility transfers, entire regions could be left without coverage for several hours:

> *I remember trying to plan with an ambulance dispatcher. I said, ‘My patient really needs to leave as soon as possible.’ Then the dispatcher, or I’m not sure who exactly, said to me, ‘Well, Doctor, are you willing to take responsibility for the fact that there are no more ambulances available in the area? [EI05_md]*

**Fig. 7.**
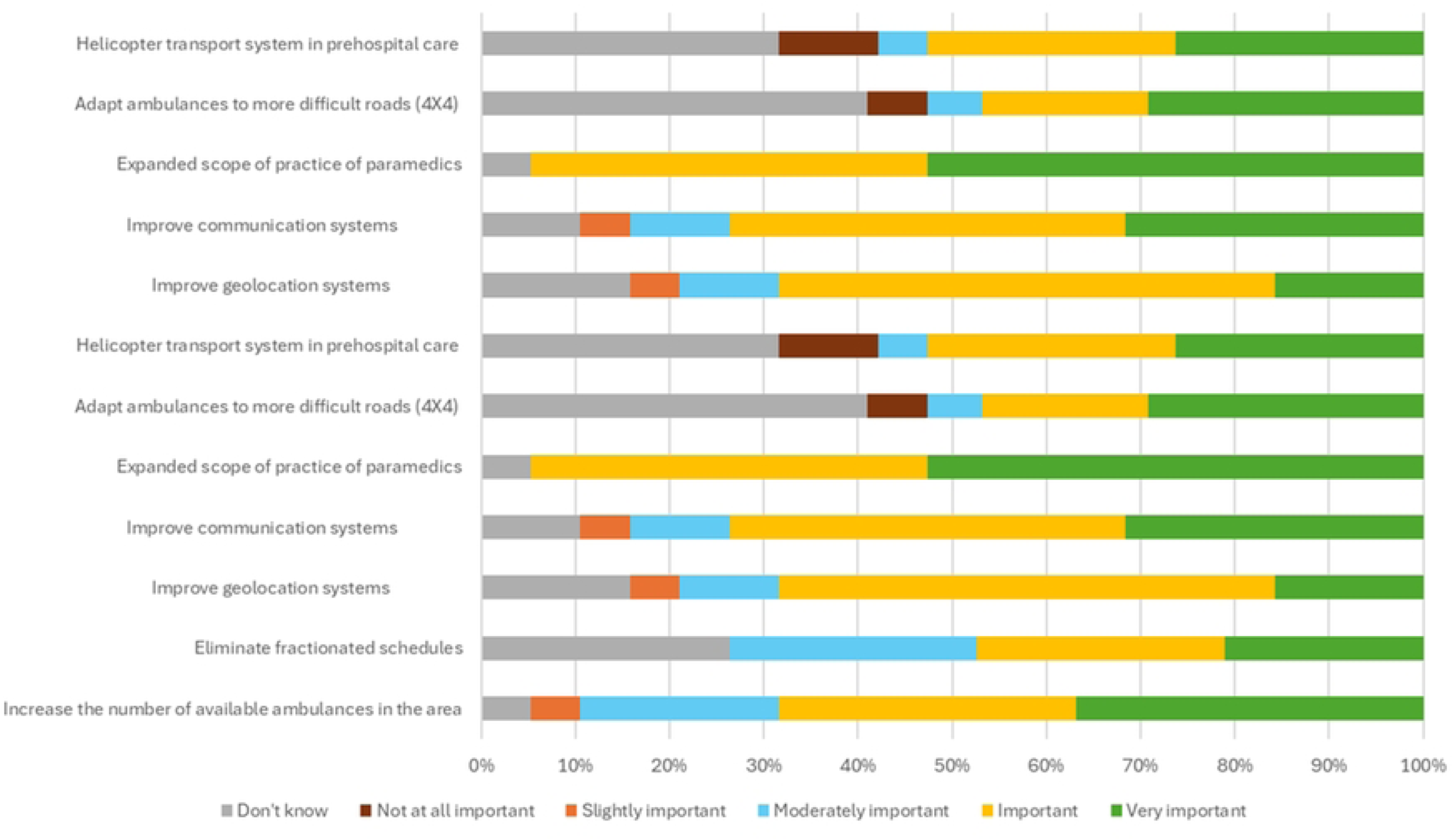
Responses (%) from champions (n = 19) to the question: “How much priority should be given to the following solutions to improve ambulance transportation?”.

### Theme 3: Access to resources

Participants emphasized persistent challenges related to infrastructure, diagnostic services, access to medical specialties, and human resources. Quantitative data and survey validation highlighted the structural nature of these barriers.

### Equipment and infrastructure

Table 6 summarizes infrastructure and equipment across rural EDs, showing gaps in areas such as resuscitation rooms and telemedicine coverage. Limited space, confidentiality, and equipment constraints, along with the need for secure, confidential telemedicine, were reported across sites. Survey respondents ranked equipment improvements and telemedicine expansion among the highest priorities (Fig 8). One practical improvement was the use of mobile equipment carts:

> *Well, recently, we made some changes to our intensive care unit. We removed cabinets and replaced them with mobile carts containing equipment. We have carts… carts that we can… Well, depending on what we need inside, there’s a cart for everything related to arterial lines, fluids, emergency medications, […] another cart for intubation equipment. These are carts that we can move around, depending on whether… if there’s an emergency in another room, we can drag all our equipment. We can move our equipment instead of moving the patient. The team appreciates this. It’s new. [EI44_inf]*

**Fig 8.**
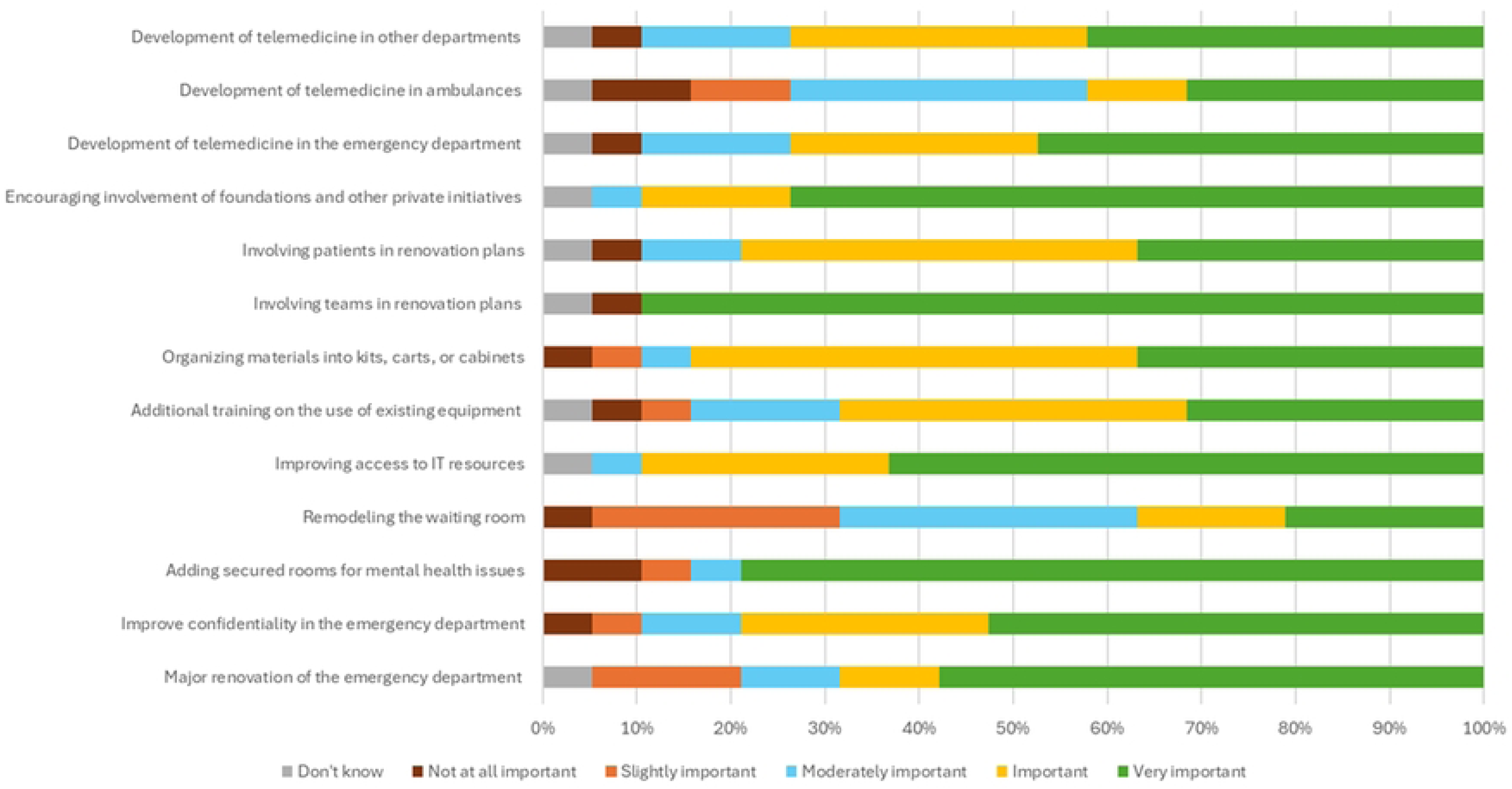
Reponses (%) from champions (n = 19) to the following two questions: “What level of priority should be given to the following aspects related to equipment and infrastructure?” and “What level of priority should be given to the development of telemedicine?”.

**Table 6.** Rural EDs with 24/7 access to diagnostic resources, 2016-2017.

| Resource | Rural EDs (n = 26) |
| --- | --- |
| Basic laboratory services | 26 (100%) |
| Basic radiology services | 26 (100%) |
| Ultrasound - portable "fast echo" device | 21 (81%) |
| Computed Tomography (CT) | 16 (62%) |
| Ultrasound - device confined to radiology rooms | 12 (46%) |
| Magnetic Resonance Imaging (MRI) | 2 (8%) |

### Diagnostic resources

Across EDs, basic laboratory and radiology services were available 24/7; many sites also had point-of-care ultrasound and CT (Table 7). A recurring issue was reliance on CT when radiology coverage was unavailable:

> *So that’s a bit frustrating too because we end up exposing our patients to much more radiation, for example, for appendicitis or cholecystitis, when ultrasound should be the primary diagnostic tool. However, since we don’t have a radiologist on-site during those times, we resort to scans. This is much more radiation-intensive, and unfortunately, it’s not recommended, but since we don’t have a radiologist, we don’t have a choice. [EI36_md]*

**Table 7.** Availability of 24/7 access to medical and paramedical specialties, 2016-2017.

| Specialty | Rural EDs (n = 26) |
| --- | --- |
| Pharmacy | 22 (85%) |
| Social Work | 16 (62%) |
| Respiratory Therapy | 19 (73%) |
| General Surgery | 18 (69%) |
| Anesthesiology | 18 (69%) |
| Internal Medicine | 12 (46%) |
| Psychiatry | 10 (38%) |
| Gynecology-Obstetrics | 6 (23%) |
| Pediatrics | 5 (19%) |
| Orthopedics | 4 (15%) |

### Medical and paramedical specialties

Access to specialties such as surgery, anesthesia, and obstetrics was uneven (Table 8). Survey respondents identified expansion of specialty coverage as a high priority (Fig 9). Participants described how prolonged gaps in anesthesia coverage threatened service continuity:

> *We find ourselves with a population threatened by service disruption and forced to suffer the consequences. There is a threat of 30 weeks without anesthesia coverage, and anesthetists are provided to us sparingly. People are informed at the last minute. Our patients end up giving birth far from their community. They cannot give birth in their community because there is no anesthetist on site. It is also important to realize that now, in the training of general practitioners and in the recommendations we make regarding emergencies, emergency physicians are not necessarily qualified to intubate a patient […] And in that situation, we find ourselves in a primary trauma center without an anesthetist, which is nonsensical. [EI71_cit]*.

**Fig. 9.**
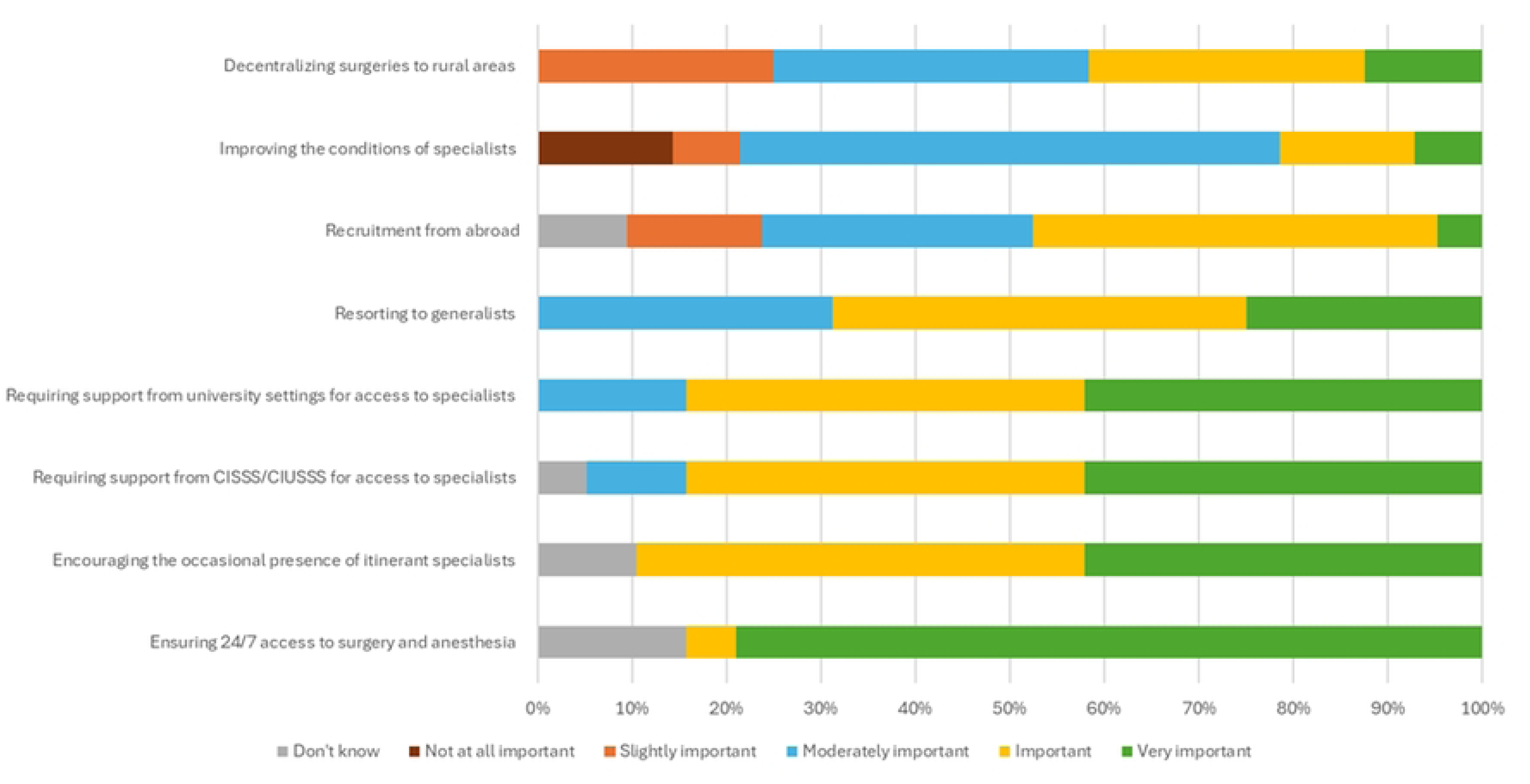
Responses (%) from champions (n = 19) to the question: “What level of priority should be given to these aspects of accessibility to medical specialties and diagnostic resources in your environment?”.

**Table 8.** Characteristics of emergency physicians at rural EDs.

| Characteristic | Rural EDs (n = 26) |
| --- | --- |
| Full-time physicians* | 4.2 ± 2.5 |
| Part-time physician** | 5.8 ± 4.4 |
| Number of MDs on shift during daytime, mean ± SD | 1.1 ± 0.3 |
| Number of MDs on shift during evening/night, mean ± SD | 1.0 ± 0.0 |
| Training as Family Physician | 92% |
| Additional training in Emergency Medicine (CCFP -EM) | 7% |
| Years of experience |  |
| 0-5 years | 40% |
| 6-10 years | 20% |
| 11-15 years | 12% |
| 16-20 years | 12% |
| Over 20 years | 16% |

\* More than 24 hours/week for a minimum of 42 weeks for clinical, academic, or administrative tasks, excluding substitute doctors. Approximate figures, as they fluctuate depending on the periods.
\*\* Those who do not qualify as "full-time," excluding substitute doctors. Approximate figures, as they fluctuate depending on the periods.

### Human resources

Staffing shortages and high turnover were recurrent themes. Table 9 and Table 10 describe characteristics of emergency physicians and nursing staff, respectively, across the 26 rural EDs. Surveyed champions also prioritized workforce stabilization (Fig 10). Participants described excessive overtime and reliance on relief staff to maintain services:

> *[The emergency] didn’t close, no. But there were about 6 periods of understaffing. […] So, the whole team absorbed it. I took on four shifts. I did 24 plus 28, I did 52 extra hours on top of my week, which was already about, maybe 60 hours. So, you know, I exceeded 100 hours last week. […] and we don’t want it to close, so we take on one more, and one more, and one more. And other colleagues also took on shifts, and everyone is a bit exhausted, you know. But no matter how much we call for help, they say, “Hold on, and don’t give up! […] We are under a lot of pressure. Because we are on so many on-call lists, we are under a lot of pressure […] Eventually, people, they stretch, stretch, stretch, they take extra shifts, and at one point, they say, “Look, I’m fed up! I’m going to the city, and I’ll practice where I want. [EI04_md]*

**Fig. 10.**
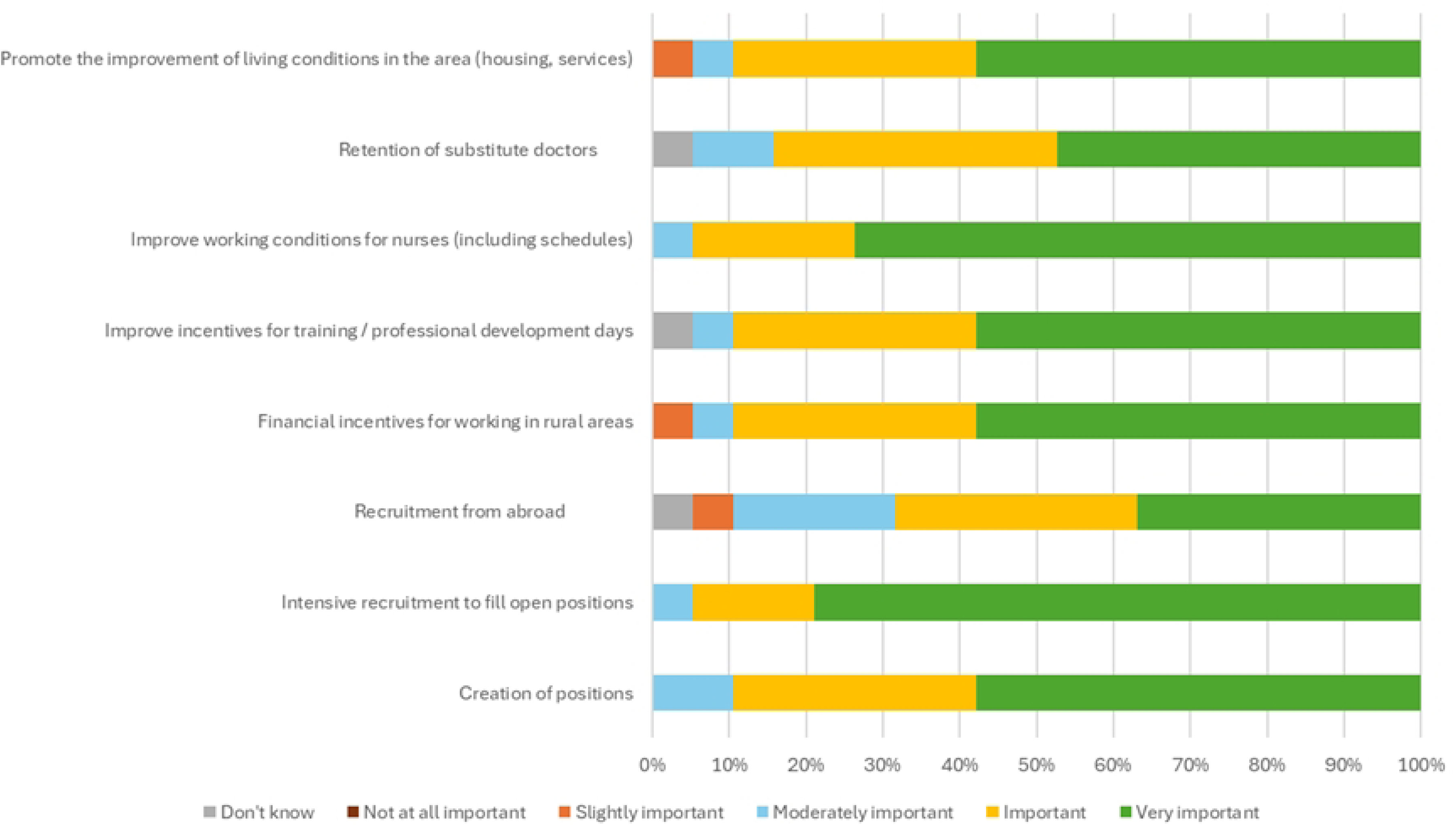
Responses (%) from champions (n = 19) to the question: “What level of priority should be given to the following aspects related to human resources?”.

**Table 9.** Characteristics of emergency nursing staff at rural EDs.

| Characteristic | Rural EDs (n = 26) |
| --- | --- |
| Full-time and part-time nurses, mean ± SD | 17.6 ± 7.8 |
| Number of nurses in ED (day), mean ± SD | 3.6 ± 1.7 |
| Number of nurses in ED (evening), mean ± SD | 3.3 ± 1.8 |
| Number of nurses in ED (night)*, mean ± SD | 2.2 ± 1.2 |
| Base nurse-to-patient ratio in Emergency Unit | 1:4.35 |
| Training as auxiliary nurse | 5% |
| Training as technician | 67% |
| Clinical training | 27% |
| Years of experience |  |
| 0-5 years | 29% |
| 6-10 years | 22% |
| 11-15 years | 17% |
| 16-20 years | 12% |
| Over 20 years | 20% |

\*Excluding on-call staff

### Theme 4: Professional practice

Participants described how rural practice requires broad versatility in low-volume contexts, often with limited on-site support. They also emphasized social proximity with patients, the importance of continuing education and simulation, and the value of strong interdisciplinary teamwork.

#### Skill maintenance

Professionals highlighted the need to maintain a wide range of skills despite infrequent exposure to complex cases. This challenge was particularly evident in low-volume settings, where staff must remain prepared for a broad scope of clinical situations despite limited hands-on opportunities:

> *You might be in the emergency department, but you’re really doing everything, everything, everything. Anything can happen, from minor surgeries to all sorts of things. I think it’s an advantage for nursing practice. You can stay up-to-date on everything, but you have to be very vigilant not to lose your skills when you don’t have many cases, to self-train, to practice with your colleagues, that can be a disadvantage. But on the positive side, it’s really diversified. It’s stimulating, depending on your perspective. [EI27_ges]*

#### Professional isolation

Several participants noted that limited specialty coverage can leave clinicians feeling professionally isolated. This sense of isolation often stems from the absence of nearby peers or specialists, which can limit opportunities for collaboration and immediate support during complex cases:

> *I would say the disadvantage we have in rural areas is that we are somewhat on our own. We don’t have other colleagues, whether in other specialties or within our own specialty, who could assist us in challenging cases? So we must be quite independent in managing our cases on-site. This is sometimes what discourages people from coming to rural areas, being alone like that. [EI16_spe]*

#### Social proximity and boundaries

Social proximity shaped everyday practice, with clinicians often navigating blurred boundaries to support access and continuity. While this proximity can foster trust and responsiveness, it also requires clinicians to navigate personal-professional boundaries in ways that are uncommon in urban settings:

> *I try to facilitate access to care for patients by giving them my cell phone number. If, for example, I operate on you, I know you inside out, I know what has been done, why would I leave you in the dark […] And there are doctors: ‘Huh, you give out your cell phone number?’ They think I get called 36,000 times a day. But that’s not the case, and it’s so much easier. [EI22_spe]*

#### Continuing education, simulation, and debriefing

Participants stressed the need for structured training, simulation, and a designated leader to coordinate quality and skills maintenance, needs that are often constrained by budgets and staffing. The absence of a dedicated clinical leader was identified as a critical gap, limiting the ability to support continuous learning and maintain standardized practices across the team:

> *Technology, and science in general, because we don’t have a dedicated clinical instructor in the emergency department. So, for that, you know, we have to do all the quality assessment, follow-ups. That would be a big plus for us. […] A clinical nurse who ensures that everyone is up to date. Not just up to date, but in terms of using, anything, like the bone drill, using thrombolysis […] Someone who would be really there for all quality follow-ups, and sharing of knowledge. [EI12_ges]*

#### Training

Training solutions that were prioritized by champions (n = 19) are shown in Fig 11. Some participants questioned whether current medical training adequately prepares clinicians for independent rural practice:

> *I realize how much mentalities, things have changed in medicine. Knowledge seems to increase very, very sharply. So there is an effort to be made to truly maintain those levels. We’re also training doctors differently. […] I’ll be blunt about it, I’m biased. But when I ask a young doctor ‘listen, when I’m told that your doctors can’t intubate a patient, it seems to me that it has always been a necessity.’ […] ‘No, well, we prefer to ask the anesthesiologist. It’s done better, and it’s more artistic.’ Well, when you’re in a rural area, I’m sorry, but the anesthesiologist might be busy in the operating room while your trauma patient arrives or your cardiac arrest patient arrives. They’re not, so to speak, trained to be ready to face it. They are ready to be in centers where there are plenty of specialists to support them. But I’m not sure they’re given training that gives them confidence to practice in rural areas. Absolutely not. [EI71_cit]*

**Fig. 11.**
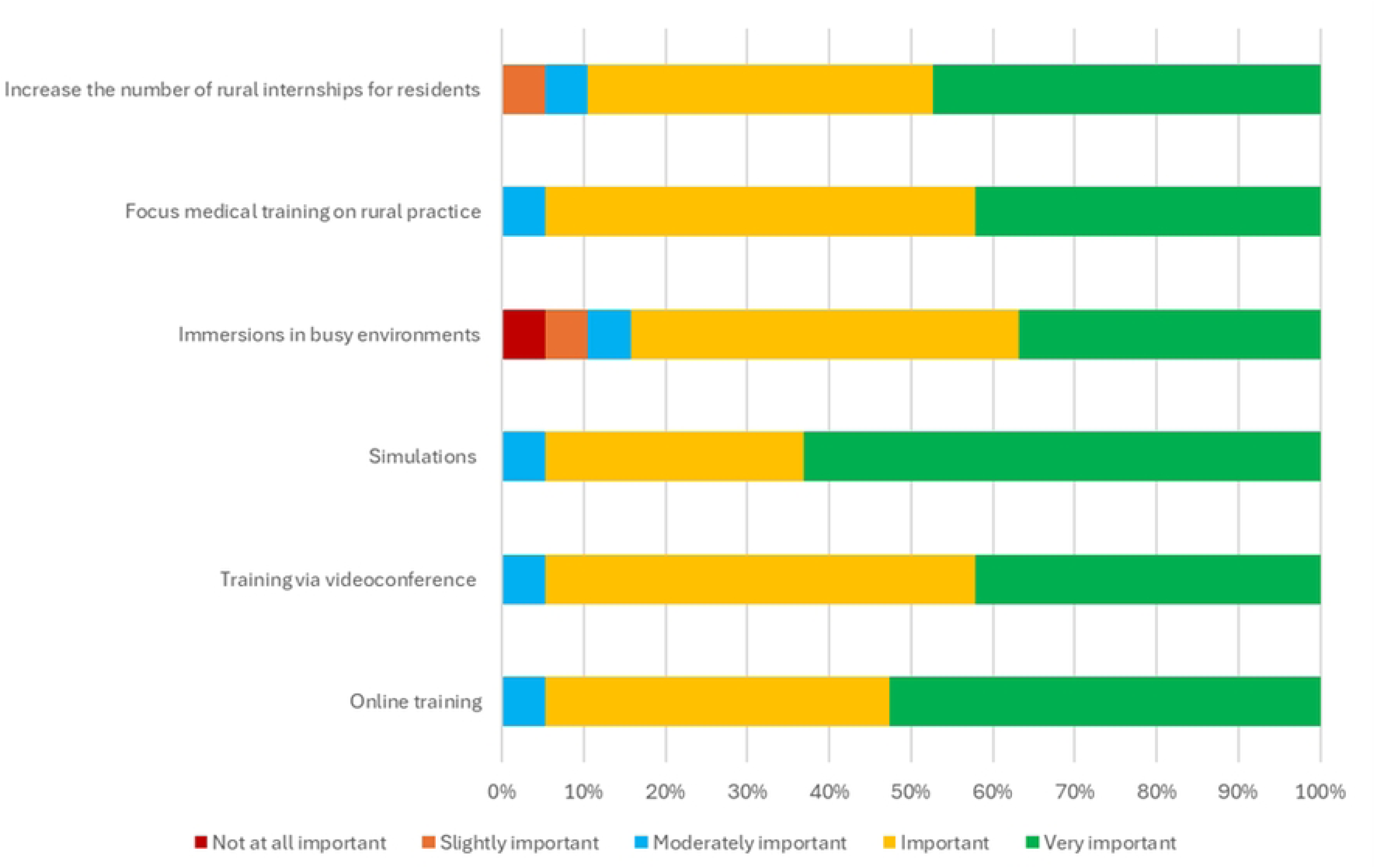
Responses (%) from champions (n = 19) to the question: “What level of priority should be given to the following training solutions?”.

#### Teamwork

Teamwork, particularly interprofessional collaboration, was highlighted by several participants as a valuable asset in rural emergency care. Due to the small size of the teams, staff members were found to be more sensitive to the effects of interprofessional conflict (Fig 12). Collegial, small-team dynamics were viewed as essential to quality care and to mitigating isolation:

> *And, you know, we’re also a small team. We know our nurses well, and we have a great dynamic with the nursing staff because it’s always the same ones, our regulars, basically. And we’re not a big team, so everyone supports each other quite well. Even when it gets overwhelming or when we have big cases, we always have a backup we can call on, and if not, well, there are other doctors in the hospital. No one ever hesitates to go help colleagues if there’s a major case coming in. [EI03_md]*.

**Fig. 12.**
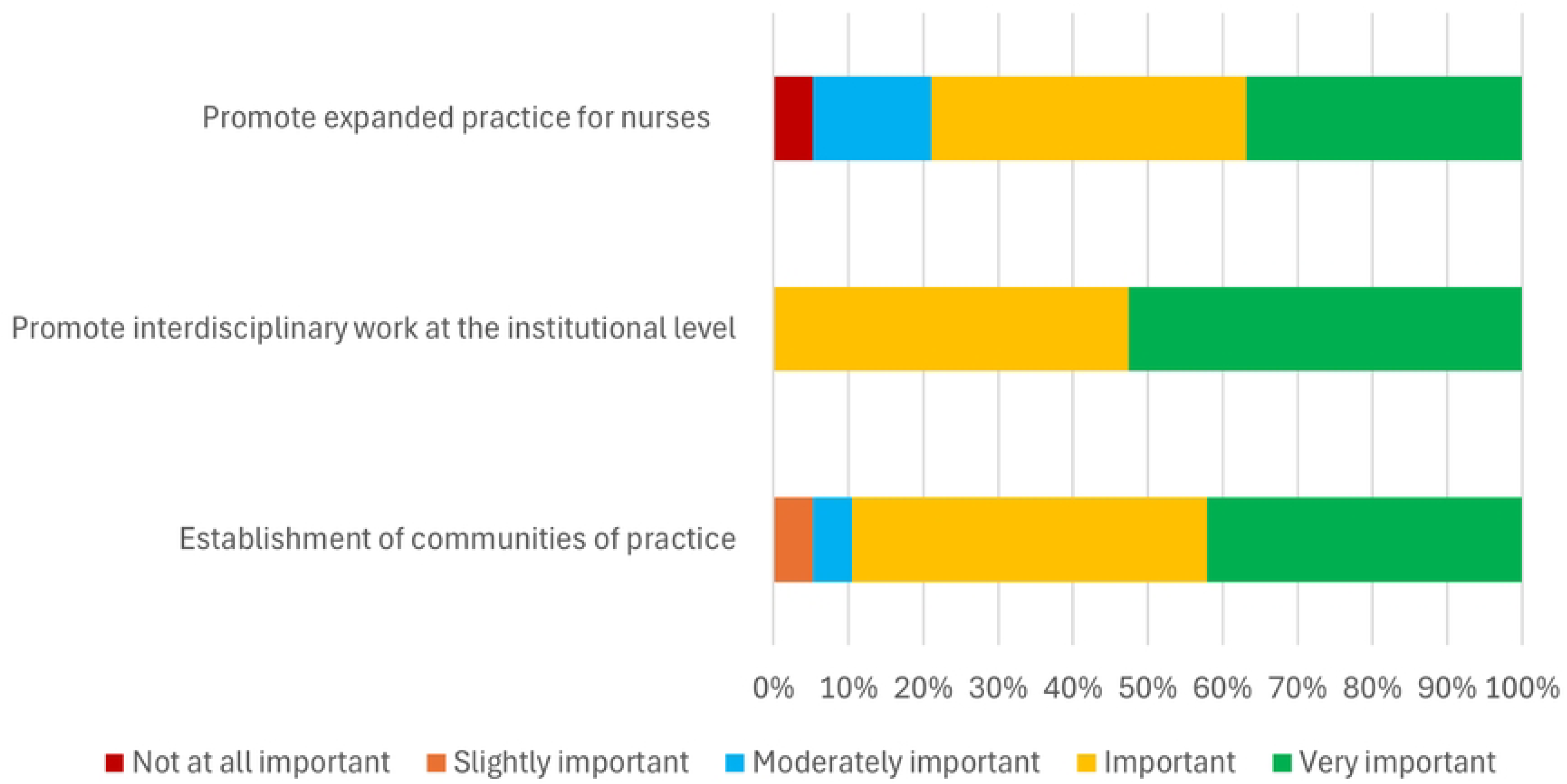
Responses (%) from champions (n = 19) to the question: “What level of priority should be given to the following training solutions?”.

## Discussion

This participatory mixed-methods study of 26 rural EDs in Québec identified four major themes shaping characteristics of ED service delivery: governance, organization of healthcare services, access to resources, and professional practice. Across these domains, participants consistently emphasized systemic challenges, particularly reduced local autonomy, workforce shortages, gaps in diagnostic and specialty services, and difficulties maintaining professional skills in low-volume contexts. Quantitative data and survey validation reinforced the structural nature of these issues, while also identifying practical solutions prioritized by local stakeholders.

### Thematic interpretation

#### Governance

Similar to other Canadian provinces, physicians in Québec are primarily remunerated through fee-for-service while other healthcare professionals are salaried [22]. While this model incentivizes service provision, it often puts rural communities with relatively few patients at a disadvantage, since volume-based metrics do not fully capture the health needs in these regions. This has become the case in Québec following the 2015 governmental reforms that created the CISSS and CIUSSS, forming a highly centralized health and social services system [23]. While the MSSS is responsible for defining strategic orientations and allocating financial resources, the CISSS/CIUSSS are charged with delivering care across vast territories, including rural zones [23]. This governance structure (combined with austerity measures, managerial reductions, and constrained public health budgets) has exacerbated regional inequities and challenged the operational flexibility of rural emergency services [11,23–25].

The findings of this study reveal that governance challenges were consistently described as systemic and far-reaching. Participants viewed the indiscriminate application of uniform budgeting models across rural regions as being poorly adapted to the realities in these areas. Many felt the administrative mergers in 2015 further centralized decision-making, leading to a reduction in local autonomy and creating a disconnect between community needs and system-level priorities. Additional bureaucratic layers delayed decision-making and discouraged local engagement. Proposed solutions included restoring localized authority through the appointment of regional emergency services managers, standardizing administrative processes within the CISSS/CIUSSS, adopting Lean management principles, and developing performance metrics tailored to rural contexts. Strengthening governance at the local level was seen as foundational to improving responsiveness, service quality, and workforce motivation.

#### Organization of healthcare services

Geographic isolation, harsh weather conditions, and limited transport infrastructure were identified as persistent barriers to timely emergency care. Inadequate access to primary care contributed to high volumes of non-urgent ED visits. Paradoxically, this demand was also perceived as necessary for maintaining ED viability in some communities. Interfacility transfers posed particular challenges, with delays compounded by limited bed availability at referral centers and insufficient ambulance resources. Suggested solutions included expanding primary care access, creating service corridors to facilitate transfers, strengthening air transport capacity, and investing in ambulance fleets and paramedic services adapted to rural conditions.

#### Access to resources

Shortages of personnel, specialty services, and diagnostic infrastructure emerged as major constraints on service quality. While telemedicine was identified as a promising support mechanism, it was only accessible in approximately half of the communities surveyed. Recruitment and retention difficulties were most acute in surgery, anesthesia, and obstetrics, threatening the sustainability of local services. Addressing these gaps will require investment in telehealth infrastructure, targeted financial incentives, improved facilities, enhanced living conditions, and coordinated recruitment strategies including international outreach and stronger university partnerships.

#### Professional practice

Maintaining diversity of clinical competencies in low-volume settings was described as a central challenge. Professional isolation, the pressures of autonomous practice, and the social proximity between providers and patients further complicated the rural practice environment. At the same time, participants emphasized the diversity of cases and the close relationships with patients as distinctive strengths of rural emergency care. Priorities for strengthening professional practice included expanding access to simulation-based training, tele-education platforms, rural–urban exchange programs, and interdisciplinary collaboration. Expanding the scope of practice for nurses was also identified as a promising, albeit complex strategy to support rural emergency services.

### Comparison with existing literature

Our findings echo prior work showing that rural EDs in Canada and internationally face barriers related to remoteness, resource scarcity, and human resource instability [26–28]. The perception of reduced autonomy following governance reforms has been described in other jurisdictions where regionalization sought to improve efficiency but unintentionally weakened local decision-making [29]. Difficulties in accessing primary care and associated ED overcrowding is also well documented in rural settings [30–32]. Similarly, interfacility transfers have been reported as critical but fragile processes in remote regions, with delays linked to both geography and workforce shortages [33,34].

Our results highlight how these systemic issues intersect: limited on-site resources increase transfers, which in turn reduce local ambulance coverage and heighten perceived vulnerability. Access to diagnostic modalities and specialty services has been identified as a key determinant of service continuity in rural hospitals [26,35]. Our study adds detail on the risks posed by prolonged gaps in anesthesia and obstetric coverage, as well as the unintended reliance on CT scans when radiology interpretation is unavailable. The professional practice findings also reinforce the literature on skill maintenance in low-volume contexts, where versatility is an asset but professional isolation poses challenges [36,37].

### Strengths and limitations

This study has several strengths. First, it encompassed all 26 eligible rural EDs in Québec, providing provincial-level coverage and ensuring a comprehensive scope. Second, its mixed-methods design enabled triangulation of quantitative data with interviews and survey validation, which enhanced both reliability and stakeholder relevance. Third, the participatory approach engaged local champions and diverse stakeholders, including citizens, in identifying challenges and solutions. Together, these features provide a rare province-wide overview of rural EDs, linking descriptive statistics with lived experiences and yielding concrete, locally validated improvement priorities.

This study also has limitations. Although all eligible rural EDs in Québec were included, the findings reflect the specific organization of the provincial health system and may not be directly generalizable to other settings. The validation survey was limited to 19 champions, which may underrepresent certain professional or community perspectives. In addition, interviews captured experiences at a single point in time, and service contexts may have evolved since data collection. As in all qualitative research, interpretation is influenced by researcher perspective; however, triangulation of data sources and a multidisciplinary analytic approach were used to mitigate this risk. Despite these limitations, the perspectives provide a rare and essential contribution to understanding and addressing rural health disparities in Canada. Moreover, within a learning health system logic and as part of a process of sharing experiences and lessons learned, they could also be highly relevant for other health systems facing similar challenges.

### Policy and practice recommendations

The findings of this study highlight the urgent need to rethink rural emergency care models. Sustainable reforms should prioritize decentralizing governance, strengthening service adaptability, and reinforcing community engagement. Local autonomy emerged as a central lever for enabling practical, region-specific solutions, while the growing geriatric and psychiatric needs of rural populations underscore the inadequacy of standardized, urban-centered models of care. Based on these findings, we propose the following eight actionable recommendations for health system leaders and policymakers:

1. **Restore and strengthen local decision-making authority.** Rural hospitals and EDs should be granted greater autonomy over staffing, service organization, and resource allocation to better align services with community needs.
2. **Formalize citizen participation in governance.** Establishing citizen and community advisory boards can ensure that local perspectives are meaningfully integrated into decision-making structures.
3. **Develop rural-sensitive performance metrics.** Indicators should reflect the realities of rural operations, rather than relying exclusively on urban-centered benchmarks.
4. **Implement tailored recruitment and retention strategies.** Targeted incentives, adapted working conditions, and customized recruitment pathways are needed to address shortages in critical areas such as anesthesia, obstetrics, and nursing.
5. **Expand access to telemedicine and virtual care.** Strategic investments in tele-radiology, tele-consultation, remote monitoring, and virtual specialist support will help bridge resource gaps across rural EDs.
6. **Strengthen interfacility transfer, prehospital services, and crisis preparedness.** Increasing ambulance availability, enhancing rural-adapted transfer protocols, expanding air medical services, and designing contingency plans for adverse weather and staffing shortages are essential to ensuring timely and safe care.
7. **Support maintenance of clinical skills and mitigate professional isolation.** Institutionalizing access to simulation training, rural–urban clinical exchanges, peer support networks, and mental health programs can sustain competencies and improve provider well-being.
8. **Monitor and report on rural–urban health equity gaps.** Transparent reporting mechanisms are needed to track disparities in access, outcomes, and resource distribution, ensuring accountability in addressing inequities.

These recommendations advocate for systemic reforms that embed local leadership, flexible service models, and community-centered innovation at the heart of rural health systems.

### Future directions and knowledge translation

The KT strategy for this study expanded significantly following additional funding from the Fonds de recherche du Québec (FRQ). Previously, a randomized trial compared three dissemination approaches: 1) a traditional webinar, 2) the written report presented in this manuscript; and 3) an ABKT intervention in the form of a 45-minute circus performance [14]. The ABKT approach generated the greatest engagement and knowledge retention, underscoring the potential of innovative, community-centered dissemination methods.

The present manuscript is an essential component of this research cycle, as it provides the first full publication of the foundational report used in the randomized trial. Its availability ensures transparency, completeness, and a reference point for future comparative KT studies. Building on the success of the ABKT initiative, additional funding has been secured to tour the 26 rural regions with interactive, community-based knowledge mobilization sessions [38]. In parallel, ongoing follow-up, local data updates, and monitoring of implemented solutions will further support the transformation of rural healthcare systems.

Finally, this work catalyzed the creation of the Living Lab Charlevoix, a rural health innovation hub dedicated to piloting and evaluating interventions that strengthen emergency care delivery [39,40]. Together, these initiatives demonstrate how evidence generated in partnership with rural communities can be mobilized to inform practice, guide policy, and inspire novel approaches to KT in emergency care.

## Conclusion

Rural EDs in Québec face persistent systemic challenges, spanning governance, service organization, resource access, and professional practice. By integrating quantitative data with stakeholder perspectives, this study identifies not only barriers but also locally endorsed solutions. The findings and recommendations presented here provide a practical framework for building a more fair and more resilient patient-centered rural health system in Québec, and potentially in other jurisdictions and countries facing similar challenges.

## Data Availability

Data cannot be shared publicly because they contain sensitive qualitative information from a small number of easily identifiable healthcare institutions and professionals in Quebec, Canada, and public release could compromise confidentiality, as required by the approving ethics committee. De-identified data may be made available upon reasonable request from Research Ethics Committee (CISSS de Chaudière-Appalaches -) to researchers who meet the criteria for access to confidential data and agree to the applicable data use conditions.

## Acknowledgements

This work was supported by the Fonds de recherche du Québec – Santé (FRQS, 22481 and 32825). The authors acknowledge Percipent Research & Consulting for providing support with language editing and thank Dr Guilherme Bresciani for his assistance with document editing and supporting the manuscript submission We sincerely thank all research participants who generously gave their time by taking part in interviews or completing questionnaires, thereby enabling a better understanding of the challenges facing rural emergency care in Quebec. A detailed list of participants is available <u>here</u>.

